# Antidepressant Use at the Threshold: using electronic health records to characterise people prescribed antidepressants around the time of dementia diagnosis

**DOI:** 10.64898/2025.12.16.25342379

**Authors:** Mohamed Heybe, Shreya Verma, Beatriz Pozuelo Moyano, Robert Stewart, Christoph Mueller, Katrina A. S. Davis

**Affiliations:** King’s College London Institute of Psychiatry, Psychology and Neuroscience, London, UK; Service of Old Age Psychiatry, Department of Psychiatry, Lausanne University Hospital (CHUV) & University of Lausanne, Prilly, Switzerland; South London and Maudsley NHS Foundation Trust, London, UK

## Abstract

**Background:** Antidepressant use is common in people with dementia. Antidepressants may be started to manage symptoms of dementia, rather than depressive and anxiety disorders. We hypothesised people prescribed antidepressants around the time of dementia diagnosis may have different characteristics from those with longstanding prescriptions.

**Methods:** We used linked primary care (Lambeth DataNet) and specialist (Clinical Record Interactive Search) data for patients with dementia in south London, UK. Antidepressant prescription was ascertained, and a new start was within one year before or after dementia diagnosis. Coded fields, a rating scale for neuropsychiatric symptoms and natural language processing of full-text were used to describe depression and anxiety.

**Results:** Of 3,713 patients with dementia, 28% were prescribed antidepressants within the year of dementia diagnosis, and 42% of these were new start prescriptions. Compared to the no antidepressant group, the new start group were more likely to be female, have vascular dementia and neuropsychiatric symptoms. Compared to the long-standing group, new start had fewer comorbidities; people from non-White ethnicities were more likely to lack documentation of depression or anxiety. Deprescribing was equally unlikely in new and long-term prescriptions (6.3% vs 5.5% per year of follow-up).

**Conclusions:** The high incidence of new prescribing, as well as the lack of deprescribing, points to unmet needs and a role for more proactive medication review. Further studies should include the clinician and patient voices to further understand how to improve non-pharmacological support for people at the threshold of dementia.

## Background

The threshold period where dementia is suspected, investigated and newly confirmed can be a confusing and distressing time. The differential diagnosis for symptoms of depression and anxiety during this time includes depressive and anxiety disorders, neuropsychiatric symptoms of dementia, and reactions to the uncertainty during the assessment process and changing social roles [1, 2]. Depression is a risk factor for dementia, and symptoms of depression often feature as a prodrome of dementia [3, 4]. Depression may also have distinct neurobiological features when appearing in someone with dementia [5, 6].

The prevalence of antidepressants in adults of all ages is approximately 23%, rising with female sex and older age [7, 8]. Some people experiencing depression and anxiety around the time of dementia diagnosis may benefit from antidepressants [9]; however, prescribing for older people, particularly those with dementia, has elevated risks [10]. Non-antidepressant strategies are considered first-line in people with dementia, including psychological treatment for depression and anxiety, personalised activities for agitation, and personalised sleep management for insomnia [1].

We aimed to characterise antidepressant prescribing around the time of dementia diagnosis, the extent to which depression and anxiety were documented, and the profiles of individuals who had recently initiated an antidepressant, especially those without a recorded diagnosis of depression or anxiety. Our findings aim to support guidance and resource planning for the clinical challenge of managing mental health symptoms in people when dementia is suspected or recently diagnosed.

## Methods

We assembled a retrospective cohort of people with dementia and identified people prescribed antidepressants in the years around diagnosis. Documentation of depression or anxiety was studied, alongside demographics, dementia features and cessation of antidepressant.

### Data Source

We used electronic health record data from the Clinical Record Interactive Search (CRIS) at South London and Maudsley NHS Foundation Trust (SLaM) [11], and a pre-established linkage to Lambeth DataNet (LDN). CRIS provides pseudonymised structured and free text data from South London and Maudsley NHS Trust (SLaM), a provider of dementia and mental health care for four London boroughs [12]. Lambeth DataNet (LDN) provides structured data about people registered with a GP in the socially and ethnically diverse London Borough of Lambeth (about 405,000 residents), one of SLaM’s catchment boroughs [13]. Natural Language Processing (NLP) has been deployed extensively in CRIS to extract a range of entities from text. Linkage includes all eligible Lambeth patients unless they have opted out of CRIS or LDN.

### Sample

We selected patients with a documented dementia diagnosis in LDN or CRIS as described previously [11] using the ICD-10 fields in CRIS and Read codes in LDN. The date of the first documented diagnosis of dementia was the index date, and eligible patients had an index date between 2009-2022, aged 65 or over, registered with a Lambeth GP and with a SLaM contact at some point from two years prior to four years post-index.

### Exposure

Antidepressant prescription was ascertained from the LDN prescription fields. The antidepressant positive group are those who were issued a prescription for antidepressants between the index date and 12 months later (the index year). We identified antidepressants through the British National Formulary [14], grouped into SSRI, tricyclic (TCA) and ‘other’ antidepressants, and matched these to the codes used by LDN (drugs + medical devices, DMD, shown in Supplementary Table S2) to search. We allocated each patient to one of three groups:

1. “New start”: prescribed antidepressant in the index year, but not in the year 12-24 months before the index date, targeting those starting in the two years between 12 months before and 12 months after the first documented dementia diagnosis.
2. “Longstanding”: prescribed antidepressant in the index year and in the year 12-24 months before index (started >1y before diagnosis).
3. “None”: not prescribed antidepressants in the index year.

A visual explanation of the group allocations is shown in Supplementary Table S1.

Using a window of observation from two years before diagnosis until four years post-diagnosis, we also noted the class of antidepressants, including the use of more than one of these in any year. Deprescription was defined as no antidepressant prescription for a complete year during follow-up. Follow-up ended after four years, at death, or if a patient left LDN according to registration data.

### Other Variables

The derivation of all variables is shown in Supplementary Table S3, and the timing is shown in Supplementary Figure S1. Demographic factors were age at diagnosis, sex, marital status, ethnicity and neighbourhood deprivation. The Charlson comorbidity index [15] (excluding dementia) from primary care codes was a measure of physical health detailed previously [11]. Dementia factors were subtype, severity at diagnosis and difficulty of activities of daily living (from closest HoNOS 65+) [16].

### Depression, anxiety and neuropsychiatric documentation

To study diagnoses and symptoms, we used:

i. Codes related to depression and anxiety: Coded mentions were primary care Read code in LDN from a list [17] or ICD-10 codes from the categories F32, F33 and F4 in CRIS at any date.
ii. Full text mentions of symptoms: Using pre-existing natural language processing (NLP) tools that identify symptoms of depression and anxiety synonyms, we assessed mentions in the CRIS text recorded in the six months either side of the index date. These are reported as being out of 21 included concepts for depression and 20 trigger words for anxiety, shown in Supplementary Table S4.
iii. A rating scale: Neuropsychiatric symptoms were derived from the clinician-completed Health of the Nation Outcome Scale (HoNOS) 65+ [16] in CRIS, the closest to the index date. One point for each problem noted (mild or above) from six (agitation, self-injury, substance use, depression, hallucinations/delusions, and other problem (which may be anxiety).

### Analyses

We carried out the following analyses:

- Antidepressant factors: We identified prescription prevalence in the index year, approximate onset, grouping the cohort into “new start”, “longstanding” and no antidepressant. We described the classes prescribed.
- Demographic and clinical factors: Characteristics were summarised using proportions or means and standard deviations for each group. Regression models were used to quantify associations between groups using linear or logistic models, as appropriate for each dependent variable.
- Depression/anxiety measures: These were summarised and compared between groups, as for demographic and clinical factors. Those with ‘new start’ of antidepressant were divided into two groups depending on whether they had a code for depression/anxiety, and clinical and depression/anxiety measures compared as previously.
- De-prescription: Deprescription was identified as above and in Supplementary Table 1. The number of deprescriptions per year of follow-up was calculated and compared between ‘new start’ and ‘longstanding’ groups using chi-squared tests.

Data was organised using R/R-studio and analysed in STATA and Excel.

The number of demographic, clinical and anxiety/depression factors was 23 for each of the two contrasts (new vs none and new vs longstanding). To avoid false discovery due to multiple comparisons, we defined significance using the Benjamini-Hochberg corrected p-value [18] less than 0.05. Appendix 3 shows the results with raw and corrected p-values. All other tables will show raw p-values, with symbols indicating the significance of the corrected p-values.

### Missing Data

Items with missing data were demographic factors of marital group and ethnicity, where an unknown group was added. In clinical features, dementia subtype contained an ‘unknown’ category for ‘unspecified’ subtype and missing. Dementia severity could only be calculated if there was an MMSE or HoNOS in the six months either side of the index date, and if not, severity was unknown.

For the other HoNOS-related outcomes, we used the closest to diagnosis, which was within six months of diagnosis for 76%. When calculating neuropsychiatric scores, we excluded those with no HoNOS ratings. Those people with no records within six months either side of the index date would score zero on depression and anxiety NLP scores if included. We were not directly able to ascertain who these were, but judged that if a person had an MMSE or HoNOS in that time-period, they should have some text for NLP. Therefore those without MMSE/HoNOD within six months of diagnosis were excluded from the NLP results. These exclusions are evident in the cohort size of Table 2 below.

**Table 1.** Sociodemographic and clinical factors for people with dementia in Lambeth DataNet, stratified by antidepressant status. Raw P-values from pairwise regression results, with significance testing from p-values corrected for multiple testing

| Characteristics | Full cohort | Antidepressant Status |  |  | New vs none |  | New vs Long |  |
| --- | --- | --- | --- | --- | --- | --- | --- | --- |
|  |  | New start | Long-standing | None | P-value | sig<sup>a</sup> | P-value | sig<sup>a</sup> |
| N | 3,713 | 441 | 599 | 2,673 |  |  |  |  |
| Socio-demographic status |  |  |  |  |  |  |  |  |
| Age at diagnosis, mean (SD) | **81.6 (7.3)** | 81.3 (7.3) | 80.7 (7.6) | 81.9 (7.1) | 0.112 |  | 0.143 |  |
| Female sex | **58.6%** | 62.6% | 68.8% | 55.6% | **0.006** | * | **0.037** | ns |
| Married | **30.6%** | 31.3% | 27.7% | 30.7% | 0.806 |  | 0.328 |  |
| Non-White Ethnicity | **38.7%** | 37.2% | 30.4% | 40.8% | 0.160 |  | **0.026** | ns |
| More deprived<sup>b</sup> | **49.0%** | 45.4% | 48.8% | 49.7% | 0.092 |  | 0.278 |  |
| Health status |  |  |  |  |  |  |  |  |
| Comorbidity score<sup>c</sup> (mean, SD) | **2.1 (1.9)** | 2.0 (1.9) | 2.5 (2.1) | 2.1 (1.9) | 0.358 |  | **<0.001** | ** |
| Dementia factors – scores from assessment closest to diagnosis |  |  |  |  |  |  |  |  |
| • Subtype |  |  |  |  |  |  |  |  |
| Alzheimer's disease | **61.8%** | 54.7% | 53.9% | 64.8% | **<0.001** | ** | 0.817 |  |
| Vascular or mixed | **18.5%** | 23.4% | 19.5% | 17.5% | **0.003** | ** | 0.136 |  |
| Other | **4.9%** | 5.7% | 7.9% | 4.0% | 0.119 |  | 0.173 |  |
| Unspecified | **14.8%** | 16.3% | 18.7% | 13.7% | 0.141 |  | 0.322 |  |
| • Dementia severity at diagnosis |  |  |  |  |  |  |  |  |
| Mild | **31.4%** | 28.8% | 30.6% | 31.4% | 0.450 |  | 0.528 |  |
| Moderate | **23.4%** | 24.7% | 23.0% | 23.4% | 0.536 |  | 0.500 |  |
| Severe | **24.7%** | 29.3% | 27.0% | 24.7% | 0.160 |  | 0.339 |  |
| Unknown | **20.5%** | 17.2% | 19.4% | 20.5% | 0.229 |  | 0.319 |  |
| • Functioning (n=3387) |  |  |  |  |  |  |  |  |
| Problems with ADLs<sup>d</sup> | **62.0%** | 66.7% | 66.0% | 60.3% | **0.015** | * | 0.817 |  |

(a) Significant association according to corrected p-value (if raw p-value <0.05): ns = p≥ 0.05, \* = p<0.05 and \*\* = p<0.01
(b) Index of Multiple Deprivations in 30% most deprived in England, based on approximate address
(c) Based on Charlson comorbidity index (with dementia not scored)
(d) Activities of Daily Living, using ADL subscale on Health of the Nation Outcome Scale (HoNOS) 65+

**Table 2.** Depression, anxiety and neuropsychiatric factors divided by antidepressant status. Raw p-values from pairwise regression for two contrasts, with significance based on p-values corrected for multiple comparisons.

| Characteristics | Full cohort | Antidepressant Status |  |  | New vs none |  | New vs Long |  |
| --- | --- | --- | --- | --- | --- | --- | --- | --- |
|  |  | New start | Long-standing | None | p-value | sig<sup>a</sup> | p-value | sig<sup>a</sup> |
| n. patients | **3,713** | 441 | 599 | 2,673 |  |  |  |  |
| Depression / Anxiety codes<sup>b</sup> ever (n=3713) |  |  |  |  |  |  |  |  |
| Depression | **32.5%** | 52.2% | 68.5% | 21.1% | **<0.001** | ** | **0.001** | ** |
| Anxiety | **21.7%** | 29.7% | 44.9% | 15.2% | **<0.001** | ** | **<0.001** | ** |
| Either | **41.9%** | 64.2% | 78.8% | 30.0% | **<0.001** | ** | **<0.001** | ** |
| Neuropsychiatric symptoms<sup>c</sup> closest to diagnosis (n=3387) |  |  |  |  |  |  |  |  |
| None | **46.9%** | 32.0% | 39.0% | 51.3% | **<0.001** | ** | 0.020 | ns |
| One or two | **44.4%** | 52.2% | 48.3% | 42.2% | **<0.001** | ** | 0.379 |  |
| Three or more | **8.7%** | 15.8% | 12.7% | 6.5% | **<0.001** | ** | 0.223 |  |
| Natural Language Processing tool score<sup>d</sup> six months either side of diagnosis (n=2934) |  |  |  |  |  |  |  |  |
| Depressive symptoms, mean (SD) | **1.9 (2.0)** | 2.8 (2.3) | 2.4 (2.5) | 1.7 (1.7) | **<0.001** | ** | 0.042 | ns |
| Anxiety symptoms, mean (SD) | **2.0 (2.0)** | 2.7 (2.3) | 2.5 (2.4) | 1.8 (1.8) | **<0.001** | ** | 0.307 |  |

## Results

### Cohort

The cohort assembly is shown in Figure 1, with 3,713 patients included in the analyses. Of the cohort, 2,539 (68%) had codes for dementia in both CRIS and LDN, while 719 (19%) had codes in CRIS only, so that 87% were definitely known to have dementia in the specialist service. Documentation of at least one MMSE or HoNOS rating within six months either side of the index date was present in 2,934 (79%). The mean follow-up time was 2.8 years (SD 2.6), with 21% alive and in LDN after four years.

**Figure 1:**
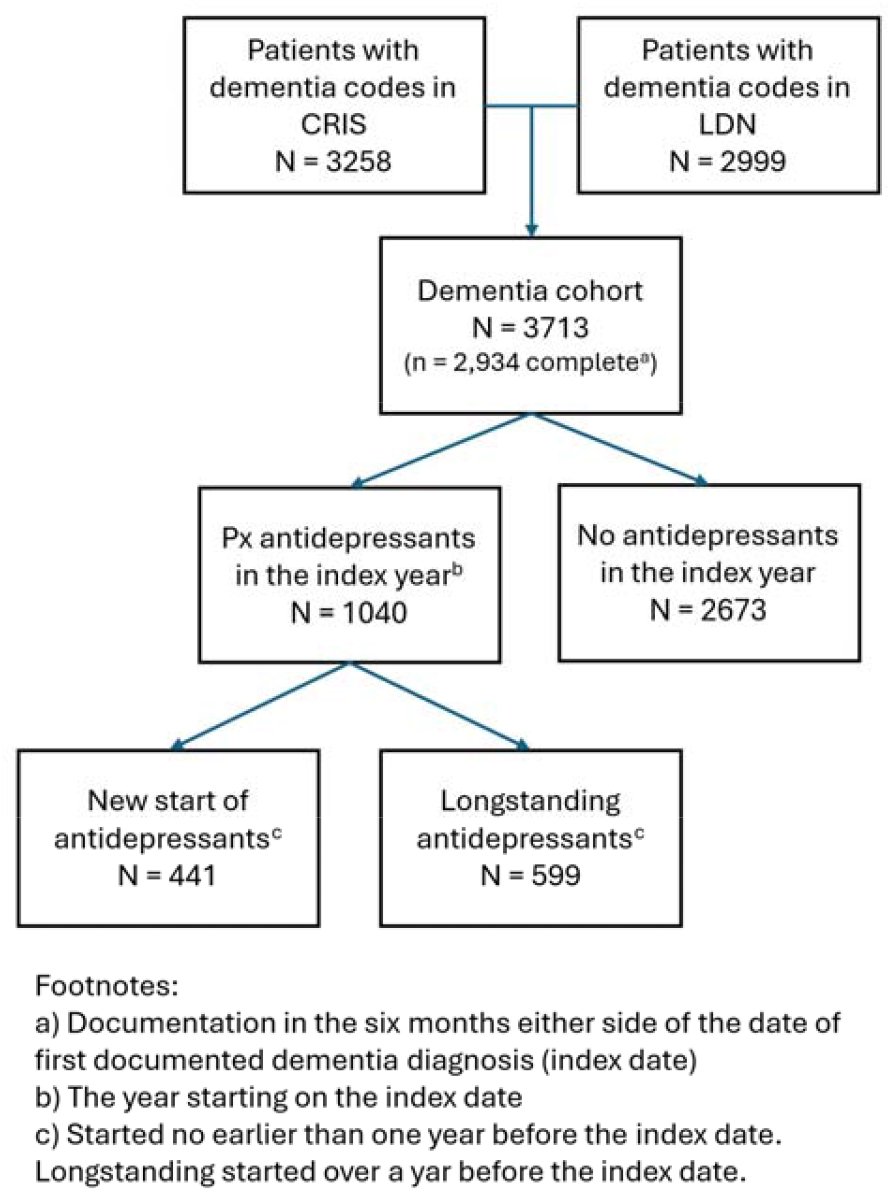
Cohort flow chart showing the formation of a dementia cohort through combining patients with a code for dementia from two sources (Clinical Records Interaction Search and Lambeth DataNet).

Characteristics of the full cohort are in Table 1. The mean age at dementia diagnosis was 82 years, 59% were female, and the most commonly recorded ethnicity was White (61%) with 6% unknown. The most common dementia subtype was Alzheimer’s disease (62%), while 31% had mild dementia at diagnosis.

### Antidepressant-related features

In the index year, there were 31,746 prescriptions for 1,040 patients (28%). For those prescribed an antidepressant in the index year, 441 (441/1040, 42%) were “new start”, and 599 were “longstanding”. Both had a mean follow-up of 2.9 years (SD 2.6).

Supplementary Table S4 shows prescribing in the index year by antidepressant class. From the 1,040, 53% received at least one prescription for an SSRI, 37% for ‘other’ antidepressants and 21% for a TCA. TCAs were prescribed for 28% (168/599) of the longstanding group and 13% (55/440) of the new start group. Twelve (12)% received a prescription for more than one of those classes. This rises to 36% receiving multiple classes over the whole observation period from two years before up to four years after diagnosis.

### Demographic and clinical factors

Table 1 describes the three antidepressant groups. When compared with no antidepressant, the ‘new start’ group showed positive associations with female gender, vascular subtype and worse ADL impairment but not with age, marital status, ethnicity, deprivation or comorbidity. When compared with longstanding antidepressant, the ‘new start’ group showed an association only with a lower comorbidity score.

### Depression/anxiety factors

Of the full cohort, 42% had at least one depression or anxiety code in GP records, and 53% had at least one recorded neuropsychiatric problem on the HoNOS. A mean of 1.9 symptoms of depression, and 2.0 anxiety-related words were identified by NLP. In Table 2, the ‘new start’ group had significantly higher scores on all of these measures compared to the no antidepressant group. Comparisons between ‘new start’ and ‘longstanding’ groups showed ‘longstanding’ had a higher prevalence of depression/anxiety codes (79% vs 64%), but after accounting for repeated testing, the groups did not differ on neuropsychiatric problems or NLP scores.

In the ‘new starters’ group, 36% did not have depression and anxiety codes in primary or specialist databases, and may have had an alternative indication. Table 3 separates those with and without formal depression or anxiety documentation. Those without such documentation were significantly more likely to come from a non-White ethnicity, and there is a suggestion they had a lower MMSE score on diagnosis. They were also less likely to score on the neuropsychiatric scale and had lower scores for NLP depression and anxiety.

**Table 3.**
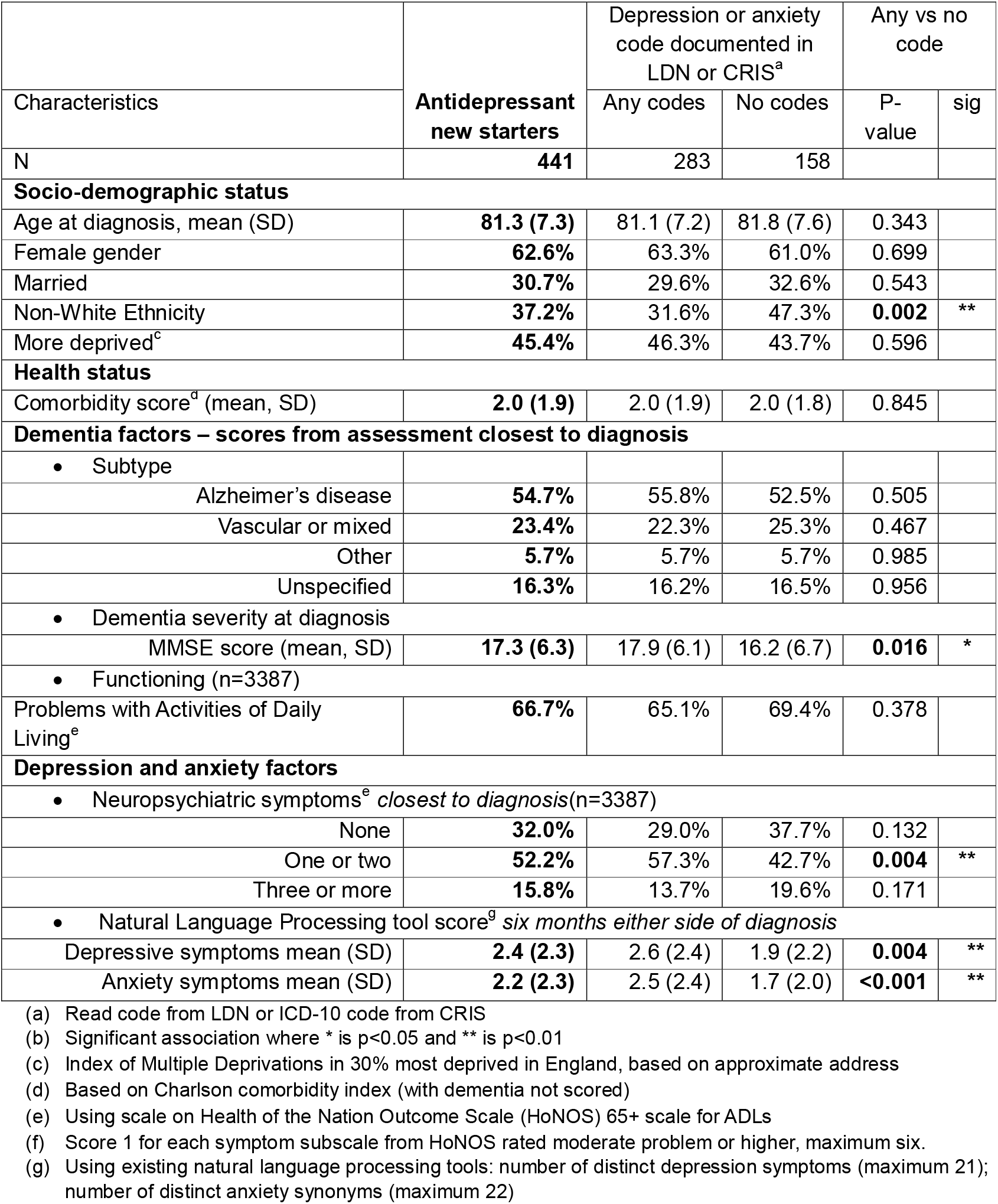
Characteristics of antidepressant new starters with separation by the documentation status of depression and anxiety. P-value and significance from pairwise regression between the documentation groups.

### De-prescription

Of the 1,040 prescribed antidepressants in the index year, 175 (17%) stopped all antidepressants for a year during follow-up. In the ‘new start’ group, 81 people (of 441) stopped antidepressants during a mean 2.9 years of follow-up: a mean of 6.3% per year of follow-up. In the ‘longstanding’ group 95 people (of 599) stopped antidepressants during a mean follow-up of 2.9 years: a mean of 5.5% per year. There was no significant difference between the groups (yearly data, p=0.557).

## Discussion

We defined a cohort of 3,713 people with dementia using routinely collected information, and defined a ‘threshold’ of dementia as one year prior to the first documented diagnosis of dementia to one year after. This is a time of suspicion and assessment, confirmation of dementia and adjustment. The prevalence of antidepressant prescription was 28% in the year of dementia diagnosis. The 441 patients who had a ‘new’ antidepressant prescription in this period accounted for 42% of those prescribed antidepressants. Compared to those without antidepressants, the ‘new’ group were more likely to be women and to have vascular dementia. To address confounding by the risk factors of depression, we also compared with longstanding antidepressant group, showing similar in demographics except lower comorbidity in the ‘new’ group. Symptom measures were higher in the’new’ group, but not after adjustment. The 36% of ‘new’ group without coded entries for depression/anxiety differed in ethnicity from those with coded entry, with lower symptom scores. During follow-up those with antidepressant prescriptions in the index year, 36% were prescribed more than one class of antidepressants and 17% (a mean of 6% per year) stopped antidepressants.

Around 30% of were prescribed TCAs, despite being cautioned in older people due to risk of cognitive decline, falls and cardiovascular issues [19, 20]. The lower use of TCAs in the ‘new’ group may mean prescribers were less keen to start TCAs in older people but may continue existing prescriptions. SSRIs were used more commonly, and though safer than TCAs, may cause QT prolongation, hyponatraemia and gastrointestinal complications in older people [21].

Vascular dementia was more common in the new start group (23%) compared to non-antidepressant group (18%), which supports the hypothesis that late-life depression may be linked to vascular pathology [22]. If depression is a marker of cerebrovascular disease, antidepressants, for which most of the evidence comes from younger adults with no vascular pathology, may be ineffective or inappropriate [5].

Depression/anxiety codes were found for 30% with no antidepressant, 79% with longstanding and 64% with ‘new’ antidepressant prescription. Therefore 36% of the ‘new’ group were without codes, similar to the 33% with no codes for depression in a UK cohort prescribed antidepressants [7]. Under-recording, a phenomenon common to dementia and mental disorders [11], will account for some of this gap. There are also alternative indications, such as pain, migraine and irritable bowel; and this may explain the higher comorbidity score in the longstanding antidepressant group. In the ‘new’ antidepressants group, some without depression/anxiety may be prescribed for distressing symptoms of dementia, such as agitation, sleep disturbances or apathy [1, 9, 23], which may account for the slightly higher neuropsychiatric score and slightly lower NLP symptom scores in these patients.

The 36% of the ‘new’ starters with no depression or anxiety codes were significantly more likely to have a non-White ethnicity. This may represent bias in healthcare. Previous work in the UK found that patients from Black ethnicities with depression were less likely to have this recorded and more likely to receive pharmacological treatment instead of psychotherapy [24]. In dementia care, people from minoritised communities have been prescribed more potentially harmful anticholinergic medication and antipsychotics on average, and fewer cognitive enhancers [25].

Where antidepressants were prescribed in the index year, they were continued in every year of follow-up in 83%. Give the prior evidence of poor efficacy in people with dementia [5, 26, 27], seen in the 36% who combined or switched antidepressant classes (a proxy for hard-to-treat depression [28]), and with no proven action of antidepressants against deterioration in dementia [10, 19], the lack of deprescribing suggests either a lack of medication reviews, a concern about stopping antidepressants, or both [29-31]. This leads to prolonged and potentially unnecessary antidepressant use.

### Strengths and limitations

The linked data sources used in this study provide a near-unique resource for these analyses in the level of detail available from multiple care providers, including comprehensive prescription data, primary care presentation coding, specialist care scales and metadata on mental health and dementia derived from NLP. This allows comparisons that would not be possible using only coded data or only one health provider. However, the data are drawn from a single area of London, so wider generalisability cannot be assumed. Routinely collected data also reflects biases in recording. For example, we previously showed that dementia was more commonly documented when cognitive enhancers were prescribed, and the same may be true for documenting depression and antidepressants.

As patient journeys are quite variable, the timings in the definitions of antidepressant start and those for symptom ascertainment were in overlapping windows. This did not allow for a consistent ordering of antidepressant start, symptom measures, and dementia diagnosis. Due to a lack of access in LDN to source text fields in the primary care record, we cannot search for more information on decisions about prescribing, and this question may be answered best by different approaches, such as qualitative research. We also miss the voices of patients and carers, which may also inform us about their beliefs. Specifically, we don’t know whether the prescription of an antidepressant means that the medication is actually taken.

## Conclusions

In a cohort from London, UK, over a quarter of people recently diagnosed with dementia were prescribed an antidepressant, and around half of these started around the time of the dementia diagnosis. There is a consensus that keeping medication burden as low as possible is preferable in people with dementia, and therefore social support and psychological treatments would be preferred over medication, although this requires the availability of appropriate services. Once antidepressants are started, they are usually continued, so there is an important role for primary and secondary care collaboration at the threshold of dementia to prevent initial prescribing, and a need for universal annual dementia medication reviews to stop any medications doing more harm than good.

## Author contributions

KASD and CM conceived the study. KASD was responsible for data curation. MH and SV conducted the statistical analysis. MH and SV wrote the first draft. KASD and CM were supervisors. All authors contributed materially to editing and approved the final version.

## Data availability

The data used in this study are available at CRIS at South London and Maudsley BRC hub, but restrictions apply for data confidentiality, see https://www.maudsleybrc.nihr.ac.uk/facilities/clinical-record-interactive-search-cris/information-for-researchers/

## Acknowledgements

This work was partially funded by the National by the National Institute for Health and Care Research (NIHR) Maudsley Biomedical Research Centre (BRC). The views expressed are those of the author(s) and not necessarily those of the NIHR or the Department of Health and Social Care. CM and BPM are members of the European Task Force for Treatment Resistant Depression in Older People.

## Funding

### Additional funding

KD is supported by the SPIN-Dementia Network+ (https://spindementianet.org/). RS is supported by the NIHR Applied Research Collaboration South London, DATAMIND HDR UK Mental Health Data Hub (MRC reference:MR/W014386) and the UK Prevention Research Partnership (Violence, Health and Society; MR-VO49879/1). CM is supported by the NIHR HealthTech Research Centre in Brain Health. BPM has received funding from the Department of Psychiatry at Lausanne University Hospital (CHUV) and a Tremplin Grant (University of Lausanne) for academic development and protected research time.

### Ethical approval

CRIS and its linkages has approval from the NRES Committee South Central Oxford C as an anonymised data source for secondary analysis ref: 18/SC/0372. This project received approval from the CRIS Oversight Committee and LDN Oversight Committee.

