## Supplementary Material for "Antidepressant Use at the Threshold: using electronic health records to characterise people prescribed antidepressants around the time of dementia diagnosis"

**Supplementary Table S1** Hypothetical scenarios with antidepressant prescribing (px) and excluded years to demonstrate the definitions of starts (which define ‘new start’ versus longstanding antidepressant groups) and stops.

**Supplementary Table S2** List of antidepressant medication used to define exposure, by class, including the SNOMED DMD codes used in LDN.

**Supplementary Table S3** Variables used in analysis

**Supplementary Table S4** Concepts and trigger words used in Natural Language Processing tools leading to depression and anxiety scores

**Supplementary Table S5** Antidepressant prescribed sample divided into new start and longstanding, and by class of antidepressant prescribed.

**Supplementary Table S6** All comparisons from tables 1 and 2 in main paper sorted by the strength of association (raw p-values) with the Benjamini-Hochberg (BH) statistic and corrected p-values

**Supplementary Figure S1**: A summary of the source and timing of variables extracted

**Supplementary Table S1**: Hypothetical scenarios with antidepressant prescribing (px) and excluded years to demonstrate the definitions of starts (which define ‘new start’ versus longstanding antidepressant groups) and stops.

|  | Years (**index** starts on date of first dementia diagnosis documentation) | | | | | | |  |  |
| --- | --- | --- | --- | --- | --- | --- | --- | --- | --- |
| ***patient scenario*** | -2 | -1 | index | +2 | +3 | | +4 | ***New / Long / None**** | ***Stop*** |
| P1 |  |  |  |  |  | |  | Long | Yes |
| P2 |  |  |  |  |  | |  | Long | No |
| P3 |  |  |  |  |  | |  | New | No |
| P4 |  |  |  |  |  | |  | New | No |
| P5 |  |  |  |  |  | |  | New | Yes |
| P6 |  |  |  |  |  | |  | None | NA |
| Key: | No prescription of antidepressant | | | | |  | |  |  |
|  | 1+ prescriptions of antidepressant | | | | |  | |  |  |
|  | Excluded (died/left cohort) | | | | |  | |  |  |

*refers to the categorisation of the cohort into groups:

New (new start) = prescribed antidepressant in the year diagnosed with dementia (index year), which started around the time of diagnosis with dementia;

Long (longstanding) = taking antidepressant in the year diagnosed with dementia and also for at least a year before dementia diagnosis;

None = not taking an antidepressant in the year diagnosed with dementia.

Antidepressant use is the prescription of any antidepressant during the index year (starting on the date of first dementia diagnosis documentation). We also studied prescribing in the year before (-1), the year two before (-2) and the three years after the index year (years +2, +3 and +4). We use the transition from a year in which any antidepressant has been prescribed to a year when no antidepressant was prescribed to define stopping antidepressant. However, we exclude years after the patient had moved or died. In table S1, patients 1 and 2 were taking antidepressants at index and also in year -2, and are labelled as having longstanding antidepressant prescriptions. Patients 3, 4 and 5 transition from no prescription to prescription by the index year, and are labelled as a ‘new start’. Both patient 1 and patient 5 transition from a year when an antidepressant was prescribed to a year when no antidepressant was prescribed, and are labelled as stopping. Conversely, patients 2, 3 and 4 are prescribed antidepressants in their last active year, and then move out of LDN or die, so this is not counted as stopping.

**Supplementary Table S2**: List of antidepressant medication used to define exposure, by class, including the SNOMED DMD codes used in LDN. Medication never prescribed in LDN returns NULL in one or more DMD columns.

| **Name** | **Medication (generic)** | **LDN DMD code** | **LDN DMD term** |
| --- | --- | --- | --- |
| **Class = Other** |  |  |  |
| Agomelatine | Agomelatine | 15499111000001100 | Agomelatine 25mg tablets |
| Amoxapine | Amoxapine | NULL | Amoxapine Tablets 25 mg |
| Amoxapine | Amoxapine | 321894004 | Amoxapine 100mg tablets |
| Amoxapine | Amoxapine | 321893005 | Amoxapine 50mg tablets |
| Brintellix | Vortiocetine | 30248111000001105 | Brintellix 10mg tablets (Lundbeck Ltd) |
| Brintellix | Vortiocetine | 30247511000001100 | Brintellix 5mg tablets (Lundbeck Ltd) |
| Cymbalta | Duloxetine | 9038411000001109 | Cymbalta 30mg gastro-resistant capsules (Eli Lilly and Company Ltd) |
| Cymbalta | Duloxetine | 9039311000001108 | Cymbalta 60mg gastro-resistant capsules (Eli Lilly and Company Ltd) |
| Duloxetine | Duloxetine | 8013111000001100 | Duloxetine 20mg gastro-resistant capsules |
| Duloxetine | Duloxetine | 417231005 | Duloxetine 30mg gastro-resistant capsules |
| Duloxetine | Duloxetine | 8013211000001106 | Duloxetine 40mg gastro-resistant capsules |
| Duloxetine | Duloxetine | 416946001 | Duloxetine 60mg gastro-resistant capsules |
| Efexor | Venlafaxine | 281211000001107 | Efexor 37.5mg tablets (Wyeth Pharmaceuticals) |
| Efexor | Venlafaxine | 628511000001107 | Efexor 50mg tablets (Wyeth Pharmaceuticals) |
| Efexor | Venlafaxine | 237411000001109 | Efexor 75mg tablets (Wyeth Pharmaceuticals) |
| Efexor | Venlafaxine | 413311000001104 | Efexor XL 150mg capsules (Upjohn UK Ltd) |
| Efexor | Venlafaxine | 30991111000001106 | Efexor XL 225mg capsules (Upjohn UK Ltd) |
| Efexor | Venlafaxine | 651811000001100 | Efexor XL 75mg capsules (Upjohn UK Ltd) |
| Isocarboxazid | Isocarboxazid | 321908006 | Isocarboxazid 10mg tablets |
| Maprotiline | Maprotiline | 321841008 | Maprotiline 10mg tablets |
| Maprotiline | Maprotiline | 321842001 | Maprotiline 25mg tablets |
| Maprotiline | Maprotiline | 321843006 | Maprotiline 50mg tablets |
| Maprotiline | Maprotiline | 321844000 | Maprotiline 75mg tablets |
| Mianserin | Mianserin | 321846003 | Mianserin 10mg tablets |
| Mianserin | Mianserin | 321847007 | Mianserin 20mg tablets |
| Mianserin | Mianserin | 321848002 | Mianserin 30mg tablets |
| Mirtazapine | Mirtazapine | 375194009 | Mirtazapine 15mg orodispersible tablets |
| Mirtazapine | Mirtazapine | 375193003 | Mirtazapine 15mg tablets |
| Mirtazapine | Mirtazapine | 9103511000001102 | Mirtazapine 15mg/ml oral solution sugar free |
| Mirtazapine | Mirtazapine | 375195005 | Mirtazapine 30mg orodispersible tablets |
| Mirtazapine | Mirtazapine | 321998002 | Mirtazapine 30mg tablets |
| Mirtazapine | Mirtazapine | 5627911000001100 | Mirtazapine 45mg orodispersible tablets |
| Mirtazapine | Mirtazapine | 375196006 | Mirtazapine 45mg tablets |
| Moclobemide | Moclobemide | 321913005 | Moclobemide 150mg tablets |
| Moclobemide | Moclobemide | 321915003 | Moclobemide 300mg tablets |
| Molipaxin | Trazodone | 75511000001108 | Molipaxin 100mg capsules (Zentiva Pharma UK Ltd) |
| Molipaxin | Trazodone | 1811000001101 | Molipaxin 150mg tablets (Zentiva Pharma UK Ltd) |
| Molipaxin | Trazodone | 369211000001100 | Molipaxin 50mg capsules (Zentiva Pharma UK Ltd) |
| Molipaxin | Trazodone | 3904511000001104 | Molipaxin 50mg/5ml oral liquid (Sanofi) |
| Molipaxin | Trazodone | 4549511000001102 | Molipaxin CR 150mg tablets (Aventis Pharma) |
| Phenelzine | Phenelzine | 321902007 | Phenelzine 15mg tablets |
| Prothiaden | Prothiaden | 208811000001101 | Prothiaden 25mg capsules (Teofarma S.r.l.) |
| Prothiaden | Prothiaden | 733111000001103 | Prothiaden 75mg tablets (Teofarma S.r.l.) |
| Reboxetine | Reboxetine | 39703311000001102 | Reboxetine 4mg tablets |
| Tranylcypromine | Tranylcypromine | 39708811000001101 | Tranylcypromine 10mg tablets |
| Trazodone | Trazodone | 321880000 | Trazadone hydrochloride* |
| Trazodone | Trazodone | 376690007 | Trazadone hydrochloride |
| Trazodone | Trazodone | 13358711000001100 | Trazadone hydrochloride |
| Trazodone | Trazodone | 35113311000001104 | Trazadone hydrochloride |
| Trazodone | Trazodone | 13358811000001108 | Trazadone hydrochloride |
| Trazodone | Trazodone | 13358911000001103 | Trazadone hydrochloride |
| Trazodone | Trazodone | 13359011000001107 | Trazadone hydrochloride |
| Trazodone | Trazodone | 321877001 | Trazadone hydrochloride |
| Trazodone | Trazodone | 13359111000001108 | Trazadone hydrochloride |
| Trazodone | Trazodone | 13359211000001102 | Trazadone hydrochloride |
| Trazodone | Trazodone | 13359311000001105 | Trazadone hydrochloride |
| Trazodone | Trazodone | 13359411000001103 | Trazadone hydrochloride |
| Trazodone | Trazodone | 13359511000001104 | Trazadone hydrochloride |
| Trazodone | Trazodone | 13359611000001100 | Trazadone hydrochloride |
| Trazodone | Trazodone | 321881001 | Trazadone hydrochloride |
| Trazodone | Trazodone | 376689003 | Trazadone hydrochloride |
| Trazodone | Trazodone | 13359711000001109 | Trazadone hydrochloride |
| Trazodone | Trazodone | 3909411000001104 | Trazadone hydrochloride |
| Trazodone | Trazodone | 13359811000001101 | Trazadone hydrochloride |
| Trazodone | Trazodone | 13359911000001106 | Trazadone hydrochloride |
| Trazodone | Trazodone | 13360011000001108 | Trazadone hydrochloride |
| Vencarm | Venlafaxine | 34180611000001109 | Vencarm XL 225mg capsules (Aspire Pharma Ltd) |
| Vencarm | Venlafaxine | 34181211000001101 | Vencarm XL 37.5mg capsules (Aspire Pharma Ltd) |
| Vencarm | Venlafaxine | 34181011000001106 | Vencarm XL 75mg capsules (Aspire Pharma Ltd) |
| Venlablue | Venlafaxine | 16421511000001100 | Venlablue XL 150mg capsules (Zentiva Pharma UK Ltd) |
| Venlablue | Venlafaxine | 23474511000001108 | Venlablue XL 37.5mg capsules (Zentiva Pharma UK Ltd) |
| Venlablue | Venlafaxine | 16421311000001106 | Venlablue XL 75mg capsules (Zentiva Pharma UK Ltd) |
| Venlafaxine | Venlafaxine | 38750411000001106 | Venlafaxine 150mg modified-release capsules |
| Venlafaxine | Venlafaxine | 14984911000001107 | Venlafaxine 150mg modified-release tablets |
| Venlafaxine | Venlafaxine | 31016011000001103 | Venlafaxine 225mg modified-release capsules |
| Venlafaxine | Venlafaxine | 15439111000001107 | Venlafaxine 225mg modified-release tablets |
| Venlafaxine | Venlafaxine | 38003311000001102 | Venlafaxine 300mg modified-release tablets |
| Venlafaxine | Venlafaxine | 23560511000001101 | Venlafaxine 37.5mg modified-release capsules |
| Venlafaxine | Venlafaxine | 18765111000001101 | Venlafaxine 37.5mg modified-release tablets |
| Venlafaxine | Venlafaxine | 39707611000001104 | Venlafaxine 37.5mg tablets |
| Venlafaxine | Venlafaxine | 14017911000001109 | Venlafaxine 37.5mg/5ml oral solution |
| Venlafaxine | Venlafaxine | 8722711000001103 | Venlafaxine 37.5mg/5ml oral suspension |
| Venlafaxine | Venlafaxine | 39707311000001109 | Venlafaxine 50mg tablets |
| Venlafaxine | Venlafaxine | 38750311000001104 | Venlafaxine 75mg modified-release capsules |
| Venlafaxine | Venlafaxine | 14985011000001107 | Venlafaxine 75mg modified-release tablets |
| Venlafaxine | Venlafaxine | 39707411000001102 | Venlafaxine 75mg tablets |
| Venlafaxine | Venlafaxine | 14018011000001106 | Venlafaxine 75mg/5ml oral solution |
| Venlafaxine | Venlafaxine | 14018111000001107 | Venlafaxine 75mg/5ml oral suspension |
| Venlalix |  | NULL | NULL |
| Vensir | Venlafaxine | 14975411000001107 | Vensir XL 150mg capsules (Morningside Healthcare Ltd) |
| Vensir | Venlafaxine | 32833611000001107 | Vensir XL 225mg capsules (Morningside Healthcare Ltd) |
| Vensir | Venlafaxine | 14975211000001108 | Vensir XL 75mg capsules (Morningside Healthcare Ltd) |
| Viloxazine |  | NULL | Viloxazine Hydrochloride Tablets 50 mg |
| Vortioxetine | Vortioxetine | 30738611000001105 | Vortioxetine 10mg tablets |
| Vortioxetine | Vortioxetine | 30738711000001101 | Vortioxetine 20mg tablets |
| Vortioxetine | Vortioxetine | 30738811000001109 | Vortioxetine 5mg tablets |
| Yentreve | Duloxetine | 8048811000001109 | Yentreve 20mg gastro-resistant capsules (Eli Lilly and Company Ltd) |
| Yentreve | Duloxetine | 8049111000001109 | Yentreve 40mg gastro-resistant capsules (Eli Lilly and Company Ltd) |
| Class = SSRI |  |  |  |
| Cipralex | Escitalopram | 3388511000001100 | Cipralex 10mg tablets (Lundbeck Ltd) |
| Cipralex | Escitalopram | 10364011000001102 | Cipralex 10mg/ml oral drops (Lundbeck Ltd) |
| Cipralex | Escitalopram | 4942511000001102 | Cipralex 20mg tablets (Lundbeck Ltd) |
| Cipralex | Escitalopram | 15453311000001105 | Cipralex 20mg/ml oral drops (Lundbeck Ltd) |
| Cipralex | Escitalopram | 7331811000001105 | Cipralex 5mg tablets (Lundbeck Ltd) |
| Citalopram | Citalopram | 321989000 | Citalopram 10mg tablets |
| Citalopram | Citalopram | 321987003 | Citalopram 20mg tablets |
| Citalopram | Citalopram | 321991008 | Citalopram 40mg tablets |
| Citalopram | Citalopram | 321994000 | Citalopram 40mg/ml oral drops sugar free |
| Citalopram | Citalopram | 407915000 | Escitalopram 10mg tablets |
| Citalopram | Citalopram | 10364811000001108 | Escitalopram 10mg/ml oral drops sugar free |
| Citalopram | Citalopram | 408067001 | Escitalopram 20mg tablets |
| Citalopram | Citalopram | 15499311000001103 | Escitalopram 20mg/ml oral drops sugar free |
| Citalopram | Citalopram | 409150001 | Escitalopram 5mg tablets |
| Escitalopram | Escitalopram | 407915000 | Escitalopram 10mg tablets |
| Escitalopram | Escitalopram | 10364811000001108 | Escitalopram 10mg/ml oral drops sugar free |
| Escitalopram | Escitalopram | 408067001 | Escitalopram 20mg tablets |
| Escitalopram | Escitalopram | 15499311000001103 | Escitalopram 20mg/ml oral drops sugar free |
| Escitalopram | Escitalopram | 409150001 | Escitalopram 5mg tablets |
| Fluoxetine | Fluoxetine | 373959000 | Fluoxetine 10mg capsules |
| Fluoxetine | Fluoxetine | 373965000 | Fluoxetine 10mg tablets |
| Fluoxetine | Fluoxetine | 321949006 | Fluoxetine 20mg capsules |
| Fluoxetine | Fluoxetine | 24407811000001108 | Fluoxetine 20mg dispersible tablets sugar free |
| Fluoxetine | Fluoxetine | 321951005 | Fluoxetine 20mg/5ml oral solution |
| Fluoxetine | Fluoxetine | 11590211000001106 | Fluoxetine 20mg/5ml oral solution sugar free |
| Fluoxetine | Fluoxetine | 32960811000001102 | Fluoxetine 30mg capsules |
| Fluoxetine | Fluoxetine | 376803002 | Fluoxetine 40mg capsules |
| Fluoxetine | Fluoxetine | 321954002 | Fluoxetine 60mg capsules |
| Fluvoxamine | Fluvoxamine | 321947008 | Fluvoxamine 100mg tablets |
| Fluvoxamine | Fluvoxamine | 321945000 | Fluvoxamine 50mg tablets |
| Lustral | Sertraline | 687611000001102 | Lustral 100mg tablets (Upjohn UK Ltd) |
| Lustral | Sertraline | 936411000001108 | Lustral 50mg tablets (Upjohn UK Ltd) |
| Paroxetine | Paroxetine | 39701911000001102 | Paroxetine 10mg tablets |
| Paroxetine | Paroxetine | 36565911000001109 | Paroxetine 10mg/5ml oral suspension sugar free |
| Paroxetine | Paroxetine | 321964006 | Paroxetine 20mg tablets |
| Paroxetine | Paroxetine | 321966008 | Paroxetine 30mg tablets |
| Paroxetine | Paroxetine | 422084009 | Paroxetine 40mg tablets |
| Prozac | Fluoxetine | 573011000001101 | Prozac 20mg capsules (Eli Lilly and Company Ltd) |
| Prozac | Fluoxetine | 695811000001109 | Prozac 20mg/5ml liquid (Eli Lilly and Company Ltd) |
| Prozac | Fluoxetine | 142311000001103 | Prozac 60mg capsules (Eli Lilly and Company Ltd) |
| Seroxat | Paroxetine | 11270611000001109 | Seroxat 10mg tablets (GlaxoSmithKline UK Ltd) |
| Seroxat | Paroxetine | 477511000001109 | Seroxat 20mg tablets (GlaxoSmithKline UK Ltd) |
| Seroxat | Paroxetine | 16811000001109 | Seroxat 20mg/10ml liquid (GlaxoSmithKline UK Ltd) |
| Seroxat | Paroxetine | 738111000001107 | Seroxat 30mg tablets (GlaxoSmithKline UK Ltd) |
| Sertraline | Sertraline | 39704011000001103 | Sertraline 100mg tablets |
| Sertraline | Sertraline | 8721711000001105 | Sertraline 100mg/5ml oral suspension |
| Sertraline | Sertraline | 39134011000001104 | Sertraline 25mg tablets |
| Sertraline | Sertraline | 13045911000001105 | Sertraline 25mg/5ml oral suspension |
| Sertraline | Sertraline | 39704111000001102 | Sertraline 50mg tablets |
| Sertraline | Sertraline | 8721911000001107 | Sertraline 50mg/5ml oral suspension |
| Class = Tricyclic |  |  |  |
| Amitriptyline | Amitriptyline | 39731711000001107 | Amitriptyline 10mg / Perphenazine 2mg tablets |
| Amitriptyline | Amitriptyline | 321745007 | Amitriptyline 10mg tablets |
| Amitriptyline | Amitriptyline | 13892411000001104 | Amitriptyline 10mg/5ml oral solution |
| Amitriptyline | Amitriptyline | 29686711000001103 | Amitriptyline 10mg/5ml oral solution sugar free |
| Amitriptyline | Amitriptyline | 8277811000001102 | Amitriptyline 10mg/5ml oral suspension |
| Amitriptyline | Amitriptyline | 39731811000001104 | Amitriptyline 25mg / Perphenazine 2mg tablets |
| Amitriptyline | Amitriptyline | 35901211000001108 | Amitriptyline 25mg modified-release capsules |
| Amitriptyline | Amitriptyline | 321746008 | Amitriptyline 25mg tablets |
| Amitriptyline | Amitriptyline | 35901411000001107 | Amitriptyline 25mg/5ml oral solution sugar free |
| Amitriptyline | Amitriptyline | 35901511000001106 | Amitriptyline 50mg modified-release capsules |
| Amitriptyline | Amitriptyline | 321747004 | Amitriptyline 50mg tablets |
| Amitriptyline | Amitriptyline | 35901611000001105 | Amitriptyline 50mg/5ml oral solution sugar free |
| Amitriptyline | Amitriptyline | NULL | Amitriptyline Hydrochloride Capsules 25 mg |
| Amitriptyline | Amitriptyline | NULL | Amitriptyline Hydrochloride Capsules 50 mg |
| Amitriptyline | Amitriptyline | NULL | Amitriptyline Hydrochloride Capsules 75 mg |
| Amitriptyline | Amitriptyline | NULL | Amitriptyline Hydrochloride Mixture Sugar Free 10 mg/5 ml |
| Amitriptyline | Amitriptyline | NULL | Amitriptyline Sr Capsules 75 mg |
| Butriptyline | Butriptyline | NULL | NULL |
| Clomipramine | Clomipramine | 321785001 | Clomipramine 10mg capsules |
| Clomipramine | Clomipramine | 321786000 | Clomipramine 25mg capsules |
| Clomipramine | Clomipramine | 4557811000001105 | Clomipramine 25mg/5ml oral solution |
| Clomipramine | Clomipramine | 321787009 | Clomipramine 50mg capsules |
| Clomipramine | Clomipramine | 36089111000001109 | Clomipramine 75mg modified-release tablets |
| Desipramine | Desipramine | NULL | Desipramine Tablets 25 mg |
| Dosulepin | Dosulepin | 321805008 | Dosulepin 25mg capsules |
| Dosulepin | Dosulepin | 8454311000001101 | Dosulepin 25mg/5ml oral solution |
| Dosulepin | Dosulepin | 321806009 | Dosulepin 75mg tablets |
| Dosulepin | Dosulepin | 8454111000001103 | Dosulepin 75mg/5ml oral solution |
| Dothiepin |  | NULL | NULL |
| Doxepin | Doxepin | 321811006 | Doxepin 10mg capsules |
| Doxepin | Doxepin | 321812004 | Doxepin 25mg capsules |
| Doxepin | Doxepin | 331643002 | Doxepin 5% cream |
| Doxepin | Doxepin | 321813009 | Doxepin 50mg capsules |
| Doxepin | Doxepin | 8484811000001101 | Doxepin 50mg/5ml oral solution |
| Doxepin | Doxepin | 321814003 | Doxepin 75mg capsules |
| Imipramine | Imipramine | 321816001 | Imipramine 10mg tablets |
| Imipramine | Imipramine | 321817005 | Imipramine 25mg tablets |
| Imipramine | Imipramine | 36046511000001104 | Imipramine 25mg/5ml oral solution |
| Imipramine | Imipramine | 16423411000001100 | Imipramine 25mg/5ml oral solution sugar free |
| Imipramine | Imipramine | NULL | Imipramine Hydrochloride Syrup 25 mg/5 ml |
| Imipramine | Imipramine | 321887002 | Trimipramine 10mg tablets |
| Imipramine | Imipramine | 321888007 | Trimipramine 25mg tablets |
| Imipramine | Imipramine | 321886006 | Trimipramine 50mg capsules |
| Iprindole |  | NULL | NULL |
| Lofepramine | Lofepramine | 39699211000001104 | Lofepramine 70mg tablets |
| Lofepramine | Lofepramine | 36040411000001100 | Lofepramine 70mg/5ml oral suspension sugar free |
| Nortriptyline | Nortriptyline | 39692911000001103 | Nortriptyline 10mg / Fluphenazine 500microgram tablets |
| Nortriptyline | Nortriptyline | 38285811000001101 | Nortriptyline 10mg capsules |
| Nortriptyline | Nortriptyline | 39701411000001105 | Nortriptyline 10mg tablets |
| Nortriptyline | Nortriptyline | 15000611000001107 | Nortriptyline 10mg/5ml oral solution |
| Nortriptyline | Nortriptyline | 37868911000001103 | Nortriptyline 10mg/5ml oral solution sugar free |
| Nortriptyline | Nortriptyline | 38285911000001106 | Nortriptyline 25mg capsules |
| Nortriptyline | Nortriptyline | 39701511000001109 | Nortriptyline 25mg tablets |
| Nortriptyline | Nortriptyline | 34034111000001108 | Nortriptyline 50mg tablets |
| Protriptyline |  | NULL | Protriptyline Hydrochloride Tablets 10 mg |
| Protriptyline |  | NULL | Protriptyline Hydrochloride Tablets 5 mg |
| Trimipramine | Trimipramine | 321887002 | Trimipramine 10mg tablets |
| Trimipramine | Trimipramine | 321888007 | Trimipramine 25mg tablets |
| Trimipramine | Trimipramine | 321886006 | Trimipramine 50mg capsules |

*terms for the generic version of trazadone were missing due to error

Supplementary Table S3 Variables used in analysis, with index being the first documented dementia diagnosis date

| **Variable** | **Source** | **Timing** | **Detail** |
| --- | --- | --- | --- |
| Basic |  |  |  |
| Sex | LDN | NA | Female / Male |
| Age (at diagnosis) | derived | index | From year of birth from LDN (approximate as year of birth, not date of birth given) |
| Marital group | CRIS | closest to index | Four categories + missing |
| Ethnicity group | LDN | most recent | Six categories + missing |
| More deprived (IMD) | LDN | closest to index | Using approximate address from LDN and IMD tables, amongst the most deprived 30% in England |
| Dementia |  |  |  |
| Subtype of dementia | CRIS/LDN | see detail | Taking CRIS most recent diagnosis if present, and LDN most often recorded subtype if no CRIS diagnosis: AD / Vascular or mixed AD and vascular dementia / Other / Unspecified |
| Dementia severity | CRIS | closest to index (within 6 months) | Severity from MMSE if there was a valid MMSE in the six month window (**valid** = has denominator >26 **and** is not 0). If no valid MMSE in window, use HONOS cognitive score if this is in six month window.  MMSE was used in 2196 cases, HoNOS was used in 800 cases, neither was available in 62 cases.  - mild=MMSE>22 or HONOS=0-2  - mod=MMSE 18-22 or HONOS=3  - severe=MMSE<18 or HONOS=4  - unknown=no MMSE or HONOS in window |
| Health |  |  |  |
| Activities of daily living problems | CRIS | closest to index | Including moderate or severe problem (score 3 or 4) |
| Charlson comorbidity index* | LDN | up until index | Up to 15 conditions (dementia not counted). Using established code lists, and original scoring. [1]  Possible score: 0-30  Actual scores: 0-13 (median 2, IQR 0-3) |
| Depression and anxiety |  |  |  |
| Neuropsychiatric symptoms | CRIS | closest to index | Number of symptoms with mild, moderate or severe problem on respective HoNOS scale (score 2, 3 or 4)  Out of 6: agitation, self-injury, substance use, depression, hallucination/delusion and other |
| Depression symptoms | CRIS | six months either side of index | Possible score: 0-20 symptoms of depression (see Supplementary Table S4)  Actual scores: 0-13 (median 0, IQR 0-2) |
| Anxiety symptoms | CRIS | six months either side of index | Possible score: 0-21 synonyms and variants (see Supplementary Table S4)  Actual scores: 0-15 (median 1, IQR 0-3) |
| Depression structured field | CRIS/LDN | ever | Binary. Mention in CRIS ICD-10 primary or secondary, Read code** for diagnosis or selected symptoms |
| Anxiety structured field | CRIS/LDN | ever | Binary. Mention in CRIS ICD-10 primary or secondary, Read code** for diagnosis or selected symptoms |
| Medication |  |  |  |
| Taking antidepressant | LDN | index year | Any antidepressant prescribed at least once in year starting on the day of first documented diagnosis of dementia (index year) |
| Start of antidepressant | LDN | year -2 | New start = started peri-dementia (taking in index year, but not taking in year -2),  Longterm = started before (taking in index year and in year -2) |
| stopped | LDN | year +1 to +3 | No antidepressant prescribed in a year before end of follow-up (4 years post index, pt died or left Lambeth GP) |
| SSRI ever | LDN | all active years |  |
| TCA ever | LDN | all active years |  |
| Other ever | LDN | all active years |  |

* Comorbidity was measured using the Charlson comorbidity index. The code lists used to define each comorbidity were derived from the Caliber project (accessible at <https://phenotypes.healthdatagateway.org/>). A spreadsheet of our codes available at DOI: 10.13140/RG.2.2.14629.42729, with definitions of the items and weights.

** See [2]

AD = Alzheimer’s disease; CRIS = Clinical Research Information Search (at South London and Maudsley); HoNOS = Health of the Nation Outcome Scales (65+); IMD = Index of Multiple Deprivations; LDN = Lambeth DataNet; MMSE = Mini-Mental State Examination; SSRI = Selective Serotonin Reuptake Inhibitor; TCA = Tricyclic Antidepressants.

Supplementary Table S4: Concepts and trigger words used in Natural Language Processing tools leading to depression and anxiety scores

| **Depressive Symptoms** | **Anxiety Synonyms** *(and Orthographic Variants)* |
| --- | --- |
| Anergia | aniexty |
| Anhedonia | anticipatory |
| Apathy | anxeity |
| Disturbed sleep | anxiety |
| Diurnal variation | anxious |
| Early morning wakening | breathless |
| Guilt | fearful |
| Helplessness | fidget |
| Hopelessness | frightened |
| Insomnia | gad |
| Low energy | jumpy |
| Poor appetite | nervous |
| Poor concentration | panic |
| Poor motivation | panick |
| Poverty of speech | restless |
| Poverty of thought | scared |
| Social withdrawal | shaky |
| Suicide ideation | tense |
| Weight loss | uneasy |
| Worthlessness | worried |
|  | worry |

Supplementary Table S5 Antidepressant prescribed in the index year divided into new start and longstanding, and by class of antidepressant prescribed. The total of the three antidepressant classes add to more than the number of people with prescription, as some are prescribed more than one.

| Taking antidepressant n | Overall | Antidepressant class | | | More than one class |
| --- | --- | --- | --- | --- | --- |
|  |  | SSRI | TCA | Other |  |
|  | **1040** | 555 | 223 | 387 | 125 |
| % of total | 100% | 53% | 21% | 37% | 12% |
| new starters n | 440 | 243 | 55 | 178 | 36 |
| % of new starters | 100% | 55% | 13% | 40% | 8% |
| longstanding n | 599 | 312 | 168 | 209 | 90 |
| % of longstanding | 100% | 52% | 28% | 35% | 15% |

Supplementary Table S6 All comparisons from tables 1 and 2 in main paper sorted by the strength of association (raw p-values) with the Benjamini-Hochberg (BH) statistic and corrected p-values

| Result number | contrast | Factor | p-value | rank | BH statistic | Corrected p-value |
| --- | --- | --- | --- | --- | --- | --- |
| 15 | None | Depression code | **0.001** | 1 | 0.041 | **0.004** |
| 44 | Long | Neuropsych =3+ | **0.001** | 2 | 0.021 | **0.004** |
| 16 | None | Anxiety code | **0.001** | 3 | 0.014 | **0.004** |
| 43 | Long | Neuropsych =1/2 | **0.001** | 4 | 0.010 | **0.004** |
| 17 | None | Either code | **0.001** | 5 | 0.008 | **0.004** |
| 41 | Long | Comorbidity index | **0.001** | 6 | 0.007 | **0.004** |
| 19 | None | Neuropsych =0 | **0.001** | 7 | 0.006 | **0.004** |
| 20 | None | Neuropsych =1/2 | **0.001** | 8 | 0.005 | **0.004** |
| 21 | None | Neuropsych =3+ | **0.001** | 9 | 0.005 | **0.004** |
| 22 | None | Depression NLP | **0.001** | 10 | 0.004 | **0.004** |
| 23 | None | Anxiety NLP | **0.001** | 11 | 0.004 | **0.004** |
| 6 | None | Alzheimer sub | **0.001** | 12 | 0.004 | **0.004** |
| 42 | Long | Neuropsych =0 | **0.001** | 13 | 0.004 | **0.004** |
| 45 | Long | Depression NLP | **0.002** | 14 | 0.007 | **0.007** |
| 7 | None | Vascular sub | **0.003** | 15 | 0.009 | **0.009** |
| 2 | None | Female | **0.006** | 16 | 0.017 | **0.017** |
| 14 | None | ADL | **0.015** | 17 | 0.041 | **0.041** |
| 38 | Long | Depression code | **0.020** | 18 | 0.051 | 0.051 |
| 27 | Long | Non-White ethnicity | **0.026** | 19 | 0.063 | 0.063 |
| 25 | Long | Female | **0.037** | 20 | 0.085 | 0.085 |
| 5 | None | More deprived | 0.092 | 21 | 0.202 | 0.202 |
| 1 | None | Age | 0.112 | 22 | 0.234 | 0.234 |
| 8 | None | Other sub | 0.119 | 23 | 0.238 | 0.238 |
| 30 | Long | Vascular sub | 0.136 | 24 | 0.261 | 0.253 |
| 9 | None | Unsp sub | 0.141 | 25 | 0.259 | 0.253 |
| 24 | Long | Age | 0.143 | 26 | 0.253 | 0.253 |
| 4 | None | Non-White ethnicity | 0.160 | 27 | 0.272 | 0.263 |
| 12 | None | Severe severity | 0.160 | 28 | 0.263 | 0.263 |
| 46 | Long | Anxiety NLP | 0.169 | 29 | 0.268 | 0.265 |
| 31 | Long | Other sub | 0.173 | 30 | 0.265 | 0.265 |
| 40 | Long | Either code | 0.223 | 31 | 0.331 | 0.329 |
| 13 | None | Unk severity | 0.229 | 32 | 0.329 | 0.329 |
| 28 | Long | More deprived | 0.278 | 33 | 0.388 | 0.388 |
| 36 | Long | Unk severity | 0.319 | 34 | 0.432 | 0.419 |
| 32 | Long | Unsp sub | 0.322 | 35 | 0.423 | 0.419 |
| 26 | Long | Married | 0.328 | 36 | 0.419 | 0.419 |
| 35 | Long | Severe severity | 0.339 | 37 | 0.421 | 0.421 |
| 18 | None | Comorbidity index | 0.358 | 38 | 0.433 | 0.433 |
| 39 | Long | Anxiety code | 0.379 | 39 | 0.447 | 0.447 |
| 10 | None | Mild severity | 0.450 | 40 | 0.518 | 0.518 |
| 34 | Long | Moderate severity | 0.500 | 41 | 0.561 | 0.561 |
| 33 | Long | Mild severity | 0.528 | 42 | 0.578 | 0.573 |
| 11 | None | Moderate severity | 0.536 | 43 | 0.573 | 0.573 |
| 3 | None | Married | 0.806 | 44 | 0.843 | 0.817 |
| 29 | Long | Alzheimer sub | 0.817 | 45 | 0.835 | 0.817 |
| 37 | Long | ADL | 0.817 | 46 | 0.817 | 0.817 |

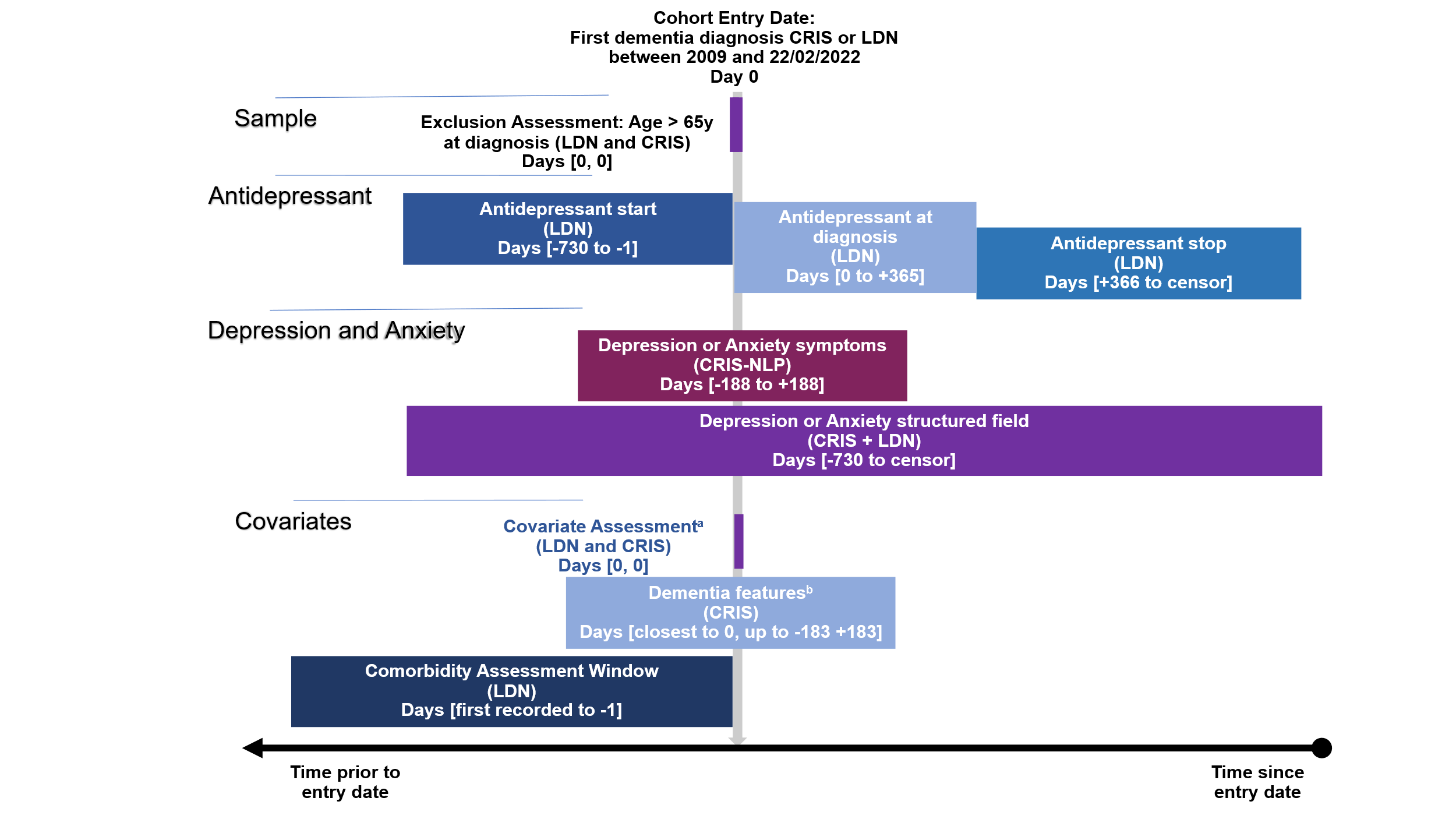

Supplementary Figure S1: A summary of the source and timing of variables extracted (based on Repeat Initiative ‘Graphical depiction of longitudinal study designs in health care databases’ [3])

a) *Sex, year of diagnosis, marital status, deprivation (area-based)*
b) *Dementia features: MMSE and HoNOS scores (inc ADL difficulty and neuropsychiatric symptoms). Also most recent dementia subtype (0 to censor)*

*Censor is until death, leaving LDN or four years post-diagnosis*
